# The Use of Conversational Agents in Self-Management: A Retrospective Analysis

**DOI:** 10.1101/2024.09.01.24312881

**Authors:** Selahattin Colakoglu, Mustafa Durmus, Zeynep Pelin Polat, Asli Yildiz, Emre Sezgin

## Abstract

**Background:** Understanding user engagement with conversational agents (CAs) in mobile health apps is crucial for improving sustained usage. We analyzed CA interactions in a mobile health app to identify usage patterns and potential barriers.

**Materials and Methods:** Retrospective data from 100,571 active users of the Albert Health app in 2022 were analyzed. Interactions with CA were categorized by demographics (gender and age), interaction type (health information, medication-related, clinical parameters, and non-clinical), and engagement method (text, voice). Descriptive statistics were used to identify trends and patterns in app usage.

**Results:** Out of the active users, 19,051 (18.9%) engaged with the CA. The majority were female (61%), with 43% aged 30-45 years and 23% older than 45 years. The analysis showed that 94.5% engaged in general health management, while 5.3% used disease-specific programs. Average usage per user was highest in cardiovascular and respiratory diseases. Interaction types varied, with health information and medication-related interactions. The varied messaging behavior suggests different user engagement levels, with some users seeking quick information and others engaging more deeply for health management. Engagement was high initially but decreased over time.

**Discussion:** This study provides insights into user interactions with a healthcare CA, highlighting a preference for general health management and diverse usage patterns. The significant number of single-session users indicates potential barriers to sustained engagement, highlighting the need for strategies to enhance user experience and retention. Future research should investigate the CA’s performance, effectiveness and extend observations to broader healthcare contexts by using large language models.

## Introduction

The growing demands on healthcare systems worldwide, increased prevalence of chronic diseases, and a push towards personalized patient care, necessitate innovative solutions.^1^ Conversational agents (CA), with their ability to automate routine interactions, offer a promising avenue to alleviate these pressures.^2^ They have the potential to impact healthcare by improving patients’ self-management and enhancing clinical practice by collecting and analyzing patient data, providing personalized feedback, and offering support for disease management and access to health information.^3^

With the increasing capabilities of eHealth and mobile health applications supported by AI, CAs have improved health communications.^4^ Literature presents that CAs have been tested in clinical settings for varying clinical phases including screening, monitoring, patient education and lifestyle coaching.^5–8^ CAs show promise in integrating into clinical practice, partnering with health professionals to monitor and assist patients, streamline clinical processes, and promote health, ultimately aiming to reduce costs, enhance operational efficiency, and elevate the quality of patient care outcomes.^9^ They have been used, as telemedicine tools, for completing routine check-ins and health-specific tasks, which supports chronic disease management, specifically with symptom tracking and medical adherence by being implemented into clinical care.^9^ Furthermore, they contribute to patient-generated health data and shared decision-making by informing HCPs about health events outside the hospital.^10,11^

Similarly, chatbots have been increasingly integrated into home care settings to enhance patient communication and care management. Recent studies demonstrated that they have been utilized for enhancing patient and family engagement while communicating medical test results^12^, for post-intervention follow-up in various healthcare interventions^13^, and for addressing health-seeking behaviors and improving patient outcomes.^2,14,15^ Additionally, they are being explored for educating chronic patients and their caregivers, demonstrating their potential to deliver personalized health information.^2,16^

However, to understand the potential benefits of CA in healthcare, it is essential to understand user interaction and adoption in a real-world setting. Current literature reported limited data on CA use and interactions in the medical field.^17^ The evidence of CA use in clinical contexts is scarce, underlining the need for understanding the characteristics of CAs.^18^ In addition, the current studies with CAs concentrated on text-based and smartphone app–delivered CAs with a focus on small case studies, suggesting further work on the feasibility, acceptability, and effectiveness of different CA formats.^1,19,20^

To fill this gap, we conducted a retrospective analysis of the usage patterns of a multimodal (voice and text) CA which is provided via a mobile application (Albert Health app) designed for health and chronic disease management.^21^ By analyzing the interaction data, we aim to understand and generate insights about the potential value and usage of CAs in healthcare and highlight the findings from CA use that promote user engagement and retention.

## Materials and Methods

For our study, we utilized a dataset composed of de-identified user interactions with the Albert CA from the year 2022. We followed a retrospective analysis, which encompassed user engagement with Albert across both iOS and Android platforms, and in two languages, English and Turkish.

### Voice-interactive mobile health assistant

Our CA (referred as “Albert” from hereon) is designed as a health assistant operating over a mobile app available in English and Turkish languages. The app is free to download and use from the Google Play Store and Apple App Store, but also available for referral by healthcare providers to invite patients to use the app. In partnership with public and private health institutions and industry, Albert is used to provide services to around 150,000 users.^22^

Albert is built using conversational development platforms and services, including Google Dialogflow and Rasa^23,24^, which provide natural language processing capabilities such as intent recognition and entity extraction. The app also uses Google Speech-to-Text (STT) and Google Text-to-Speech (TTS), to enable voice input and output and Speech Synthesis Mark-up Language (SSML) to enhance Albert’s natural and expressive responses. The main service that includes these NLP (Rasa and ML frameworks provided by Dialogflow) and STT (Google Speech) models which communicates with the mobile application and makes the necessary guidance, is called *AI Engine*. It sends the audio bytes in stream format to the speech-to-text models and forwards the resulting text to *NLP models*. See figure 1 and “An example of user interaction” section for flow of user engagement and explanation (*Fig. 1*).

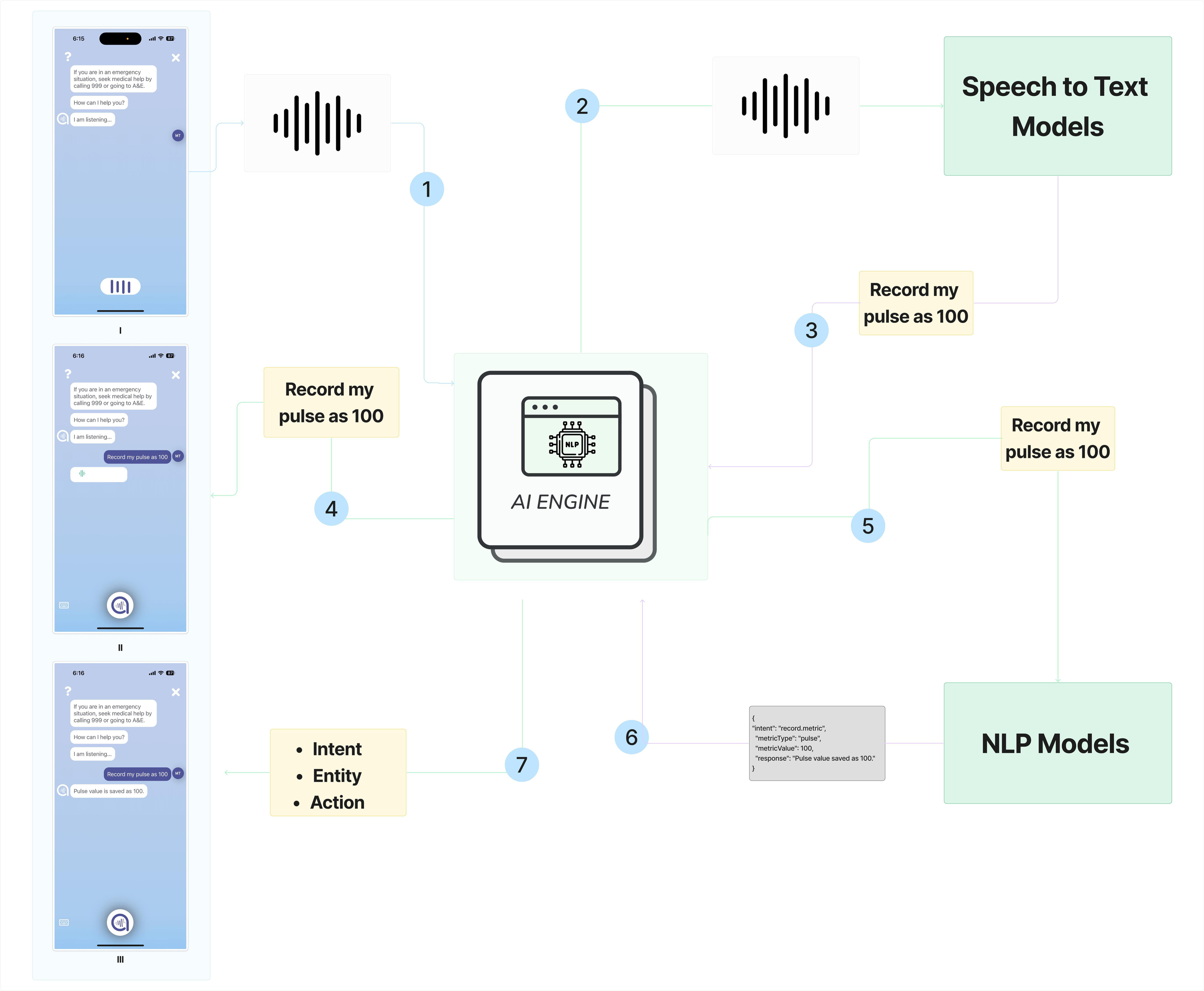

### An example of user interaction

1. The user initiates an interaction with the mobile application by using a voice command, such as "Record my pulse as 100." This audio input is captured by the app’s interface.
2. The audio data is then sent to the Google Cloud Speech API, a service that can process spoken language into text. The API uses advanced machine learning models to accurately transcribe spoken words into written text.
3. The transcribed text, which reads "Record my pulse as 100," is then sent from the Google Cloud Speech API back to the application’s processing system.
4. The application displays the transcribed text within its graphical user interface.
5. This transcribed text is sent to the AI engine. The AI engine passes text to the NLP model to analyze the structure and semantics of the sentence to extract meaningful information.
6. The NLP model identifies the intent, entities, and proposed actions from the user’s command. Intent refers to the user’s objective (e.g., recording health data), entities are the relevant data points or parameters involved (e.g., the pulse rate), and actions are the operations the system needs to perform (e.g., log the pulse rate in the user’s health records).
7. The extracted intent, entities, and actions are then processed by conversational AI frameworks such as RASA or Dialogflow. These frameworks manage the conversational flow and ensure that the user’s command is executed correctly. They are capable of handling complex dialogues, maintaining context, and managing the conversation state.
8. In the mobile application, once the intent and entities are confirmed, the app would proceed to record the user’s pulse rate as 100 in their personal health record. The graphical interface serves as a confirmation and error-checking mechanism, allowing users to verify that their voice command was understood and executed correctly.

The app is used for general health management or disease-specific self-management purposes. Disease-specific self-management programs (DMPs) are designed to address a range of health conditions across various medical specialties, including endocrinology (e.g., Diabetes Mellitus, Growth Disorders), cardiology (e.g., Hypertension, Heart Failure), nutrition (e.g., dietary management for chronic illnesses), neurology (e.g., Multiple Sclerosis), and respiratory medicine (e.g., Asthma) Each DMP was customized by considering the needs of specific patient groups and some have a disease-specific CA which lets users get information about their chronic diseases and record some disease-specific parameters. Non-DMPs are designed for general health support and promote healthy behavior without focusing on any certain disease.

### Intents

Intents were divided into four groups: Access to health information, treatment adherence, clinical parameter tracking, and others. All conversations about health information seeking, such as FAQs, tips, pharmacies, appointments, medication info, and asking for advice, were included in the “Health Information” group. All conversations, such as medication reminders, were included in the “Medication adherence” group. All conversations that help clinical decision making, such as Blood Pressure, Blood Glucose Level, Body Mass Index, and Disease Journal, were included in the “Clinical decision support” group. Fallback (responses when the agent doesn’t understand a query), smalltalk (casual conversations), technical support, and guidance were included in the "Others" group (*Table 1*).

**Table 1.** Intent groups, explanations and examples.

| Intent Category | Definition | Conversational examples |  |
| --- | --- | --- | --- |
|  |  | Non-DMP<br>(General health management) | DMP (Disease specific) |
| Health Information | Interactions about general health advice, or medical knowledge that could be relevant to both general well-being and specific health conditions.<br>Example:<br>- Ask health-related questions,<br>- Show pharmacy on duty,<br>- Get a telehealth appointment | User: What's an ideal body fat percentage for women?<br>CA: Ideal body fat percentages vary by individual and depend on factors like age and fitness level. Generally, for women, a body fat percentage of 21-24% is considered healthy. Athletes might have lower percentages, around 14-20%, while levels above 32% might be considered higher than optimal for health. It's important to focus on overall well-being and consult with a healthcare professional for personalized advice. | User: I'm not sure if my inhaler technique is right.<br>CA: Ensuring the correct technique is crucial for your treatment. Could you tell me the name of your inhaler?<br>User: Ventolin HFA.<br>CA: I'll send you a video for Ventolin HFA inhalers. If you have any questions after watching, I'm here to help! |
| Clinical Parameters | The process of tracking, recording, and reviewing health metrics such as blood pressure, blood glucose level, or other clinical data<br>Example:<br>-Record blood pressure or glucose level,<br>- Measure BMI,<br>-Record a symptom diary | User: Can you calculate my BMI?<br>CA: Absolutely. Please provide me with your height and weight.<br>User: I'm 175 cm and weigh 70 kilos.<br>CA: Based on that, your BMI is 22.9, which is within the healthy weight range. If you want to schedule an online meeting with a dietitian, please let me know. | User: I want to log my blood pressure.<br>CA: Please tell me your latest reading.<br>User: It's 13 over 8.<br>CA: I've recorded your blood pressure as 130/80 mmHg. |
| Medication - Related | Actions related to the management and understanding of medications, supplements, or treatments, applicable to routine health maintenance or condition-specific management.<br>Example:<br>- Record medication,<br>- Mark medication as taken,<br>- Show daily medications | User: Can you show my daily medication schedule?<br>CA: Certainly.<br>- 08:00g: Vitamin D, 1000 IU with breakfast.<br>- 18:00: Omega-3 supplement with dinner. | User: I need to record my new medication.<br>CA: Of course. Please tell me the name of the medication.<br>User: Metformin.<br>CA: Got it. I've added Metformin to your medication list. What time will you be taking it? |
| Non-clinical | Casual interactions and system guidance to ensure a comfortable and supportive user experience, these address general inquiries and navigate uncertainties.<br>Example:<br>-Fallback,<br>-Smalltalk,<br>-Technical Support & Guidance | User: [Attempts to speak, but the speech is unclear and not transcribed correctly]<br>CA : I'm sorry, I didn't catch that. Could you please repeat what you just said?<br>User: [Attempts to speak again, but the speech is still unclear and not transcribed correctly]<br>CA: It seems we're having trouble understanding your message. I apologize for the inconvenience. If you're experiencing an urgent issue, it's important to contact a healthcare professional directly or dial emergency services. Remember, this platform is not a substitute for professional medical advice, diagnosis, or treatment. Please try to rephrase your concern or reach out to your healthcare provider for immediate assistance. |  |

### Recruitment and study setting

We included users who engage with Albert across both iOS and Android platforms, and in two languages, English and Turkish in 2022. To recruit patients for these programs, three main methods are used: patients download the app themselves from the store, healthcare professionals invite patients during hospital visits, and insurance companies recommend the app as part of their disease management initiatives. Our inclusion criteria is users from Turkey interacted with the conversational agent in 2022 at least one-time. We excluded those who entered an invalid birthdate (> 2022, < 1900).

During the installation, users were requested to provide consent for research purposes. This consent process was designed to ensure that participants were fully informed about the nature, scope, and implications of data collection and subsequent analysis activities associated with our research. Following that, users completed an onboarding survey about their demographics and disease-related conditions the first time entering the app. This study received approval from the Biruni University Ethics Committee (Approval number: 2023/83-18). During the use period, users received a one-time reminder notification on the 8th day after the installation.

### Data privacy

In adherence to privacy standards, all data procured for the study were removed from personal identifiers, such as names and contact details. To further safeguard participant confidentiality, each user was assigned a distinctive identifier. The application, purpose-built for health and chronic disease self-management, autonomously recorded user interactions with the CA.

Encryption protocols were employed to ensure the privacy of the data. Throughout the analysis, user anonymity was preserved via the use of the aforementioned unique identifiers. Our analytical focus was restricted to pre-existing data, with no additional data collection undertaken.

### Data analysis

We analyzed the data from 19,016 users who completed the tutorial of CA and used the chat function within the app between January 1, 2022, and December 31, 2022. Users were included as active users if they entered the app at least once in 2022, and chat users if they used Albert in 2022. The data included user demographics, disease management program (DMP) participation, and detailed user engagement data such as the number and types of interactions with Albert. We conducted descriptive analysis and analyzed the frequency and mode of user-initiated interactions (i.e., touch or voice) and the distribution of user interactions over time.

## Results

Out of the 100,571 active users of Albert Health app between January 1, 2022, and December 31, 2022, 19,016 (18.9%) completed the tutorial and interacted with Albert.

### Demographics

In the demographic profile of the study’s participants, 61% (7,717) of the users identified as female, while 39% (4,898) were male, and gender was not reported for 34% (6,436) of users. Age-wise, 42% (7,918) did not report their age. Among those who did, 8% (876) were between 0-15 years, 25% (2,731) were between 15-30 years, the largest age group was 30-45 years at 45% (4,979), and 23% (2,547) were 45 years or older (*Table 2*).

**Table 2:** User Demographics.

| Demographics | Number of Users | Percentage |
| --- | --- | --- |
| **Gender** |  |  |
| Female | 7706 | 61% |
| Male | 4881 | 39% |
| Not reported | 6429 | 34% |
| **Age** |  |  |
| Not indicated | 7909 | 42% |
| 0-15<sup>1</sup> | 873 | 8% |
| 15-30 | 2722 | 25% |
| 30-45 | 4970 | 45% |
| 45 or more | 2542 | 23% |
| **DMP users- non-specific to any health condition** | 18001 | **94.66%** |
| **DMP users- with a specified health condition** | 1015 | **5.34%** |
| **DMP-Nutrition** | 598 | **3.14%** |
| **DMP-Neurology** | 229 | **1.2%** |
| **DMP-Respiratory** | 53 | **0.27%** |
| **DMP-Oncology** | 46 | **0.24%** |
| **DMP-Cardiology** | 31 | **0.18%** |
| **DMP-Endocrinology** | 32 | **0.16%** |

<sup>1</sup> Users indicated their ages between 0-15 are anticipated to be parents or caregivers creating an account on behalf of their children.

### Health condition-specific findings

Within the study categorization, users engaged in health condition-specific programs—including cardiology, oncology, neurology, and endocrinology—represented 5.34% (1,015 users), with a total of 5.6% (5,369 interactions), averaging 5.3 interactions per user. General health management was utilized by 94.66% (18,001 users), accounting for 89.5% of interactions, with an average of 4.9 interactions per user (*Table 3*). Regarding the types of interactions, health information queries constituted the majority (27%, n=24,921), followed by clinical parameters at 20% (n=18,378), and technical support queries at 18% (n=16,521). Casual small talk comprised 14% (n=12,949), while fallback queries were at 12% (n=11,825). The least engaged category was medication-related, with 9% (n=8,889) (*Table 3*).

**Table 3:** Engagement Rates and Program.

| Category | Users (%) | Interactions (%) | Average Usage per User |
| --- | --- | --- | --- |
| General Health Management | 18,001 (94.6%) | 88,452 (94.3%) | 4.9 |
| Disease-specific program | 1,015 (5.3%) | 5,369 (5.7%) | 5.3 |
| Subgroup: Respiratory Disease | 53 (0.2%) | 360Interactions | 6.8 |
| Subgroup: Cardiovascular Disease | 31 (0.2%) | 341 interactions | 11 |

### Engagement and interaction

Users interacted with Albert most frequently (47%) for getting health information, followed by medication-related topics (36%) and clinical parameters (17%) in the clinical context (*Table 4*).

**Table 4:** Interaction Categories and Initiation Methods.

| Category | Number of Interactions (Frequency) | Initiated by Screen - Touch | Initiated by Voice |
| --- | --- | --- | --- |
| Health Information | 24,921 (27%) | 56% | 44% |
| Clinical Parameters | 18,378 (20%) | 53% | 47% |
| Medication-Related | 8,889 (9%) | 57% | 43% |
| Technical Support, Guidance & Other unsolicited queries | 16,521 (18%) | 58% | 42% |
| Smalltalk | 12,949 (14%) | 12% | 88% |
| Fallback | 11,825 (12%) | 16% | 84% |
| *Total* | 94,530 100%) | 48% | 52% |

Of the interactions related to getting health information, 56% were initiated by touch (tapping the app on the screen), and 44% by voice. Interactions related to medication tracking and clinical parameters, frequencies were also similar to getting health information, as mostly initiated via touchscreen. Among “non-clinical” types of interactions, voice-based interactions were 2 times higher than touch-based interactions (68% vs. 32%). Out of those, 11,231 users engaged for just one session (single session users) and 7,785 users with two or more sessions (multi-session users). Similarly, 10,268 users sent 1 or 2 chat messages, while 8,748 users sent two or more messages. In the first 50 days after registration, there was a peak on the first day with the most significant number of sessions made and a smaller peak on around day 10 (after reminder), and then a decreasing trend was observed. Our observation was intentionally limited to 50 days, as we did not observe any changes for the following days (*Fig. 2*).

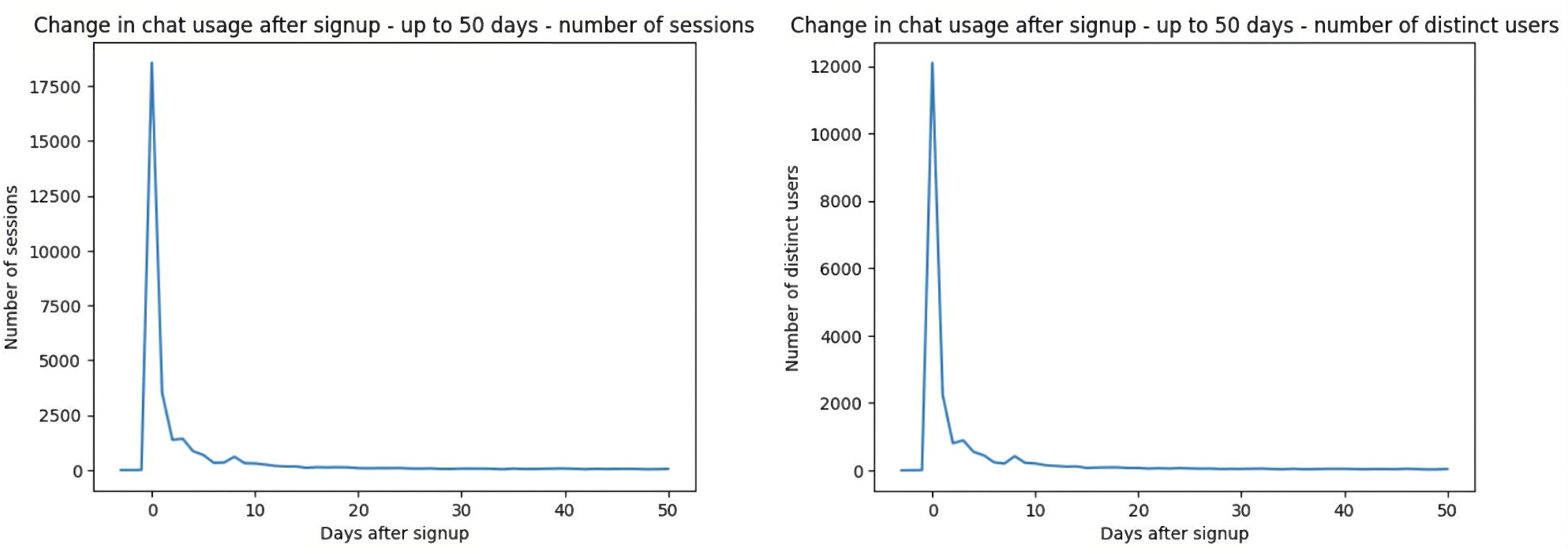

## Discussion

Our study underscores the increasing role of CA in the healthcare domain and prevalence of its use by patient populations, particularly in facilitating chronic disease management and general health monitoring.

The analysis of user demographics revealed a dominant engagement among female users, a trend that may reflect gender differences in health-seeking behavior and the proactive management of health via mobile technology.^25^ The less frequent disclosure of age could suggest privacy concerns or a lower perceived relevance of age information in app interaction.^26,27^ However, the reported age distribution were similar with the CA studies that most of the participants were young and middle aged adults^3^, indicating a similar user profile among CA users for CDM. Intriguingly, the segment reporting on behalf of young children underscores the app’s indirect reach to pediatric health management.

The preference for general health management programs over disease-specific ones could indicate a user tendency toward preventive health measures^3^ and the convenience of accessing a broad spectrum of health resources in a single platform.^28^ The considerable engagement in category interactions, which encapsulate a diverse range of topics beyond health information, medication, and clinical parameters, including small talk (casual conversations), technical support, and guidance. That indicates the versatility of the CA in addressing a wide array of user needs and questions might be preferable.^29^ In addition, Fallback instances demonstrated that the most common issue the message has not been truly understood by CA. This can be due to two reasons: first, the user is not synced with the assistant and terminating session before the message complete; second, the failure of speech-to-text in cases where the user mentions medical terminology or infrequently used drug names (for example, "Hodgkin’s lymphoma"). Other cases include the user not saying anything meaningful or using slang. Such instances have also been observed in the literature with earlier CA encounters in healthcare, failure of understanding statement or context.^30,31^ This shows there is a room for improvement in healthcare context when developing such tools and there is need for continuous evaluation of CAs. In addition, large language models can improve natural language understanding in cases where medical terminology was not captured.

The interaction patterns present a significant insight. While screen touch is the predominant mode of initiating interactions, the substantial use of voice commands, especially in the small talk and fallback (+80%) categories, signals user adaptability and the potential for more hands-free, accessible app navigation in certain contexts. One interesting note on interactions with voice-activated interfaces is that there is a notable inclination towards anthropomorphizing the technology, with users frequently initiating conversations with social inquiries and greetings, treating the system as a conversational partner^32^. Additionally, the voice interface serves as a primary channel for technical support requests, diverging from clinical queries. These trends could inform enhancements in the app’s design to better accommodate and encourage voice interactions, possibly reflecting user convenience or the necessity for accessibility in certain scenarios.^1,33^

### Retention

The fact that a significant number of users (n=11,251) engaged with the app for only a single session suggests there may be barriers to sustained use, such as user expectations not being met, usability issues, or a lack of perceived value in continued interaction.^34–36^

Contrastingly, 7,785 users who engaged in two or more sessions represent a more committed segment, potentially indicative of a group finding ongoing utility in the app’s features. This dichotomy presents an opportunity for targeted user experience improvements and personalization to convert single-session users into regular users. Similarly, the messaging behavior—where 10,268 users sent only 1 or 2 messages, while 8,748 users had a higher engagement with two or more messages—highlights varying levels of interaction depth. It could reflect different user needs or satisfaction levels with the information and support provided.^37^ For instance, users sending fewer messages might be utilizing the app for quick, specific information, while those with more messages could be leveraging it for more comprehensive health management. Understanding the drivers behind these different usage patterns could inform the development of features towards both brief interactions and more detailed health discussions, optimizing the CA to serve a spectrum of user engagement preferences.^1,37^

Finally, the initial surge in user engagement immediately post-registration, with a second, albeit smaller peak around day 10, aligns with typical digital engagement patterns.^38^ This might indicate an initial curiosity or a critical period where users explore the app’s features, followed by a decline as the novelty wears off. Identifying factors that contribute to sustained engagement beyond this period could be pivotal in increasing the long-term adoption and therapeutic impact of health-related CAs. Strategies to retain users could include personalized health insights, reminders for health tracking, or gamification of health tasks to maintain a consistent user-app interaction trajectory.^39,40^

### Limitations

The study is subject to several limitations. Firstly, the data are observational and based on user interactions within a single application, limiting the generalizability of the findings to other CAs or health apps. Secondly, the reliance on self-reported data and the substantial proportion of users not disclosing their gender or age could introduce bias and affect the accuracy of demographic insights. Additionally, the high rate of single-session users might indicate a selection bias, as the data primarily represents users who chose to engage with the app, potentially excluding those who discontinued use after initial download. The study also lacks qualitative data on user satisfaction and reasons for discontinuation, which could provide deeper insights into user behavior and preferences. Finally, the retrospective nature of the analysis limits the ability to establish causal relationships between app features and user engagement patterns, necessitating further research to explore these dynamics comprehensively.

## Conclusions

The study provides insights into user engagement with a healthcare CA, indicating a predominant use for general health management and a diverse interaction pattern, which can be complementary to current telemedicine applications. These findings underscore the need for personalized and user-friendly features to address varying user needs and preferences, ultimately enhancing sustained engagement and effectiveness of health CAs. Further research is recommended to explore the causal factors influencing app usage and to generalize these findings to other healthcare applications.

## Data Availability

All data produced in the present study are not available due to ethical committee approval

## Contributions

S. Colakoglu: Conceptualization (lead); writing – original draft (lead); formal analysis (lead); writing – review and editing (equal). M. Durmus: Software (lead); Data curation (lead); writing – review and editing (equal). Z.P. Polat: Writing – review and editing (equal). A. Yildiz: Writing – original draft (supporting); Writing – review and editing (supporting). E. Sezgin: Supervision (lead); Conceptualization (supporting); Writing – review and editing (supporting).

